# Prevalence and risk factors of chronic kidney disease in a populated area with highly moving populations

**DOI:** 10.1101/2020.10.19.20215004

**Authors:** Mohammad Khajedaluee, Sanaz Ahmadi Ghezeldasht, Arman Mosavat, Reza Hekmat, Seyed Abdolrahim Rezaee, Tahereh Hassannia

## Abstract

**Background:** Chronic kidney disease (CKD) is a major public health problem worldwide. Prevalence and associated risk factors of CKD was evaluated in the greater Mashhad, a highly populated pilgrimage city, in 2018 on 2,976 subjects.

**Methods:** This study was carried out in the greater Mashhad, a highly populated pilgrimage city, in 2018 on 2,976 subjects. For each participant a standard questionnaire, a physical examination and clinical history were completed. Then biochemical and hematologic tests for kidney function were performed.

**Results:** Obesity was observed more frequently in subjects with CKD, thus BMI was positively correlated with the prevalence of CKD (*p*<0.001). Moderately reduced GFR was found in 17.6% and 7.1%, and severely reduce GFR at 0.7% and 0.5%, of males and females, respectively (*p*<0.001).Drug abuse also showed a strong association with CKD (*p*=0.004), but smoking did not. Using univariate and multivariate logistic regression of decreased estimated GFR<60 showed that age (OR=1.06), gender (OR=2.14), diabetes (OR=1.07), hypertension (OR=1.39) and drug usage (OR=3.29) were risk factors for CKD; BMI was not. The same statistics showed that only age (OR=1.02), diabetes (OR=2.61) and hypertension (OR=1.16) were risk factors for albuminuria. The prevalence of hypertension (22.1%) was a risk factor for CKD, and vice versa. BMI and drug abuse were also risk factors for hypertension, but not smoking.

**Conclusion:** These findings demonstrated that progression of CKD and hypertension in any population should be considered in the context of changes in human behaviours, etiology, disease severity, co-occurring diseases, addiction and priority of therapy over prevention.

## INTRODUCTION

Chronic kidney disease (CKD) is becoming a major problem worldwide for health authorities because of its trend progression and its association with cardiovascular diseases.It is among the main causes of death that has radically increased in the last 30 years, due to aging, urbanization, and emerging diabetes, and hypertension, lack of access to adequate healthcare,herbal medication and incomplete knowledge of its pathogenesis.Nowadays, researchershave demonstrated that diabetes and hypertension are the two main causes of kidney disease in the developed world.^1^

The most important danger for patients and health authorities are, firstly,the risk of the disease progressing toward end-stage renal disease (ESRD), a statusrequiringhemo or peritoneal dialysis or kidney transplantation,which are costly and debilitating. Secondly, CKD is ahigh-risk factor for cardiovascular diseases (CVD).^2-4^By understanding the risk factors and pathogenesis of CKD in its early stages, it is possible to reduce the occurrence of CVD and ESRD.Because of the nature of the early stages of CKD, it is not primarily diagnosed until its progression toward kidney damage has commenced; therefore, more studies and attention are needed to reduce the development and progression of renal failure and its adverse consequences. Even serious kidney damage can be delayed through early diagnosis and CKD therapy.^5,6^

According to 2002 KDOQI Guidelines, definition of CKD is a glomerular filtration rate (GFR) below 60 mL/min/1.73 m2 for three months or more,irrespective of the cause and with or without evidence of kidney damage.^7^GFR is assessed by serum creatinine usingthe Cockcroft and Gault (CG) and Modification of Diet in Renal Disease (MDRD) equations for adults.^7,8^

In the United States, a national survey, demonstrated that the prevalence of CKD had an increased trend from 10% in 1988–1994 to 13.1% in 1999–2004.^9^Studies also showed a high prevalence of CKD in Japan’s general population, around 18.7%.^10,11^In Asia, Europe and Australia, similar trends of the high prevalence of CKD have been reported.^12^

However, less adjusted and reliable data has been reported from developing countries, particularly in the Middle East.In Iran, a high prevalence of CKDhas been observed among people over age 20. In another study, the prevalence of CKD was calculated by MDRD for the general population of Tehran, and was found to be 18.9% (95% CI 18.2, 20.6).^13^

For appropriate management of kidney diseases and prevention or slowing of their progression, it is pivotal to assessaccurate prevalence rates at particular stages in different geographic regions. Therefore, in this study,the prevalence and the associated risk factors of CKD were evaluated in a large, population-based study of Mashhad in Iran. The general population of Mashhad is 3 million, and, because it is a pilgrimage city, its moving population is around 20-30 million per year.

## METHODS

### Study population and sample size

All participants completed a written informed consent, which was approved by the Biomedical Ethics Committee of Mashhad University of Medical Sciences. The study was performed on the general population of the greater Mashhad between May and September 2018.

The list of all households under coverage of the city’s 20 district health centres (primary health and treatment services) was used for sampling.A random sample of the households was taken, stratified by healthcare centre to achieve a distribution similar to the original population. In each household, one person above the age of 16 was recruited so that the study included the both genders as well as appropriate percentiles for ages according to the 2011 census in each district.There were 2,976 full participants in this study. Of this sample, 1,045 were male (35.1%) and 1,931 (64.9%) were female.

Prior to the beginning of the study a four-day official workshop was coordinated by a nephrologist, a socio-medical specialist and a biochemist for one physician from each of the district health centres to discuss the kidney failure signs and symptoms, staging, hypertension and the priority of precise data collection.Then, medical examinationswere performed on participants and histories and demographic and anthropometric data were taken by these physicians. Blood pressure was assessed by a mercury sphygmomanometer. Height and weightweremeasured using aSeca217 and 707, respectively. Body mass index (BMI) was also recorded. The inclusion criteria including;age > 16 year, resident of the district more than 1 yearand filling informed consent. Furthermore, the exclusion criteria were dialysis treatment, any transplantation, malignancy, HIV infection diagnosis and pregnancy or pregnancy within the past 6 months. The flowchart 1 shows all the relevant of study variables.

### Sample collection and serological assay

Blood samples were taken between 8-10am after a 10-12 hours overnight fast according to the protocol of Mashhad’s ACECR-Central Medical Lab.Analyses of blood biochemical agents such as fasting glucose, total cholesterol (TC) and triglyceride (TG), creatinine and BUN concentrations were carried out at the Central Laboratory of ACECR using aSelectra 2 autoanalyser (Vital Scientific, Netherlands).

Urinary factors, includingcolour, appearance, pH, protein, ketones,Hb, bilirubin,urobilinogen, blood, glucose, nitrite, bacteria, parasites, crystal, mucus, yeast, candida, cast,WBC, RBC and Epithelial cell were examined.

### Definition of chronic kidney disease

A simplified MDRD equation^14^was used for GFR estimation in this study as follows: GFR (mL/min/1.73 m2) = 186.3 * (serum creatinine)-1.154 * (age)-0.203 * (0.742 if female) * (1.21 if African American).

### Statistical analysis

Data were analysed using SPSS version 16(SPSS Inc., Chicago, IL, USA). The analyses were carried out using descriptive statistical indexes including standard deviation, mean and a confidence interval at 95%.

For data analysis, the distributions of the variables were estimated using the Kolmogorov– Smirnov test.For variables with normal distribution, the parametric test was used, and, for others, non-parametric tests were used. For qualitative variables, a chi-square test was used.For non-normal distributed variables, Man Whitney and Kruskal Wallis tests were used.For quantitative variables,*t*-student and one-way ANOVA tests were used. Univariate and multivariate logistic regression tests were employed to discover risk factors for CKD. The resultswere significant if *p*≤ 0.05.

## RESULTS

### Socio-demographic data

Out of the 2,976-full-participant study population, 1,045 of them were male (35.1%) and 1,931 (64.9%) were female. A total of 22.1% participants (20% of women and 52.9% of men) had hypertension, 8.8% (6.8% women and 12.6% men) had diabetes, and 39% had a history of chronic diseases. Finally, 4.86% of women and 14.9 % of men had a history of smoking and 1.8% of participants were drug users(Table 1).

**TABLE 1.** The characteristics of study population according to the gender.

| Character | Male (n=1044) | Female (n=1930) | Total (n=2974) | p-value |
| --- | --- | --- | --- | --- |
| Age(y) | 46.1(15.75) | 42.12(13.89) | 43.52(14.69) | <0.001 |
| **Age group** |  |  |  |  |
| <30 | 19.41 | 24.8 | 22.9 | <0.001 |
| 30-39 | 16.5 | 18.4 | 17.8 |  |
| 40-59 | 43.4 | 44.9 | 44.4 |  |
| 60-69 | 11.5 | 8.8 | 9.7 |  |
| >70 | 9.1 | 3.2 | 5.3 |  |
| Body mass index <sup>a</sup> | 25.72(4.15) | 27.15(5.03) | 26.65(4.79) | <0.001 |
| **Body mass index (classification %)** |  |  |  |  |
| <18 | 3.8 | 3.3 | 3.4 | <0.001 |
| 18-25 | 39.8 | 31.9 | 34.7 |  |
| 25-30 | 42.5 | 38.3 | 39.8 |  |
| 30-35 | 11.4 | 19.8 | 16.9 |  |
| >35 | 2.5 | 6.7 | 5.3 |  |
| Smoking% | 14.92 | 4.86 | 8.40 | <0.001 |
| Drug abusing% | 3.8 | 0.7 | 1.8 | <0.001 |
| Diabetes, diagnosed% | 11.7 | 6.5 | 8.3 | <0.001 |
| Hypertension, diagnosed% | 25.9 | 20.0 | 22.1 | <0.001 |
| History of chronic disease% | 39.2 | 38.9 | 39 | 0.89 |
| Urine albumin, mg/dL | 21.8(2.63)*<br>5.3(1-12)** | 18.4(1.72)<br>5.7(1-12) | 19.59(1.45)<br>5.47(1-12) | 0.25 |
| Urine creatinine, mg/dL | 142.82(3.04)<br>130(90.35-183.65) | 125.8(2.14)<br>115(73-164.5) | 131.7(1.76)<br>122(77.8-172.7) | <0.001 |
| Serum creatinine, mg/dL | 1.23(0.009)<br>1.2(1.1-1.3) | 1.04(0.007)<br>1(0.9-1.1) | 1.11(0.006)<br>1.11(1-1.2) | <0.001 |
| Estimated GFR,<br>71.29mL/min/1.73m<sup>2</sup> | 71.29(0.62)<br>69.58(61.6-80.4) | 84.84(0.53)<br>84.26(84.2-96.8) | 80.15(0.43)<br>78.8(66.6-92.2) | <0.001 |
| <sup>a</sup>Calculated as weight in kilograms divided by height in meters squared. |  |  |  |  |
| *mean (SD) |  |  |  |  |
| ** median (25-75 percentiles) |  |  |  |  |

As Table 1 shows, the mean age of the population was 43.52 ± 14. 69 years, ranging from 16-60, with an age distribution profound in the 40-59 level and older, in which CKD seems to be more common.A total of 36% of the subjects were under the age of 40, and 60% were over the age of 40. This indicates that age is a risk factor for CKD, with CKD being more frequent in older participants (*p*<0.001). The mean BMI of 25-30 was more frequent in this study (39.8%),and is in the range of being overweight. Taken together, 22.2% of the subjects were in the range of obesity, 39.8% were in range of being overweight, and 38.1%fell in the normal range. Around 62 % of the population fell in the range of being overweight,which are risk factors for CKD and CVD. BMI has a strong association with the prevalence of CKD (*p*<0.001), and obesity in this population was more frequent where CKD was more prevalent. While smoking was not significant factor for CKD occurrence, drug abuse had a strong association with CKD (*p*=0.004).

Furthermore, although the mean albumin in urine in men (21.8±2.63) and women (18.4±1.72) was not statistically significant, the mean urine creatinine in men (142.82±3.04) and women (125.8±2.14) was significant (*p*<0.001). The mean concentration of serum creatinine in men was 1.23 mg/dL and in women was 1.04 mg/dL.This difference was significant (*p*<0.001). The mean estimated GFR, 71.29mL/min/1.73m^2^ in men was 71.29 and in women was 84.84,a statistically significant difference (*p*<0.001). Taken together, the indices for renal function in females was generally better than in men (Table 1).

### Kidney function and albuminuria

According to the GFR, mL/min/1.73 m^2^, about 12.1% of men and 40.3% of women had a GFR over 90 and in range of normal; 69% of men and 52% women had a GFR 60-89, in range of mildly reduced kidney function. Moreover, around 17.6% of men and 7.1% of women had moderately reduced GFR, and 0.7% of men and 0.5% of women had severely reduced GFR. The differences between men and women in these categories were significant(*p*<0.001). Table 2demonstrates kidney function according to GFR, mL/min/1.73 m^2^ (A) and albuminuria (B).

**TABLE 2.**
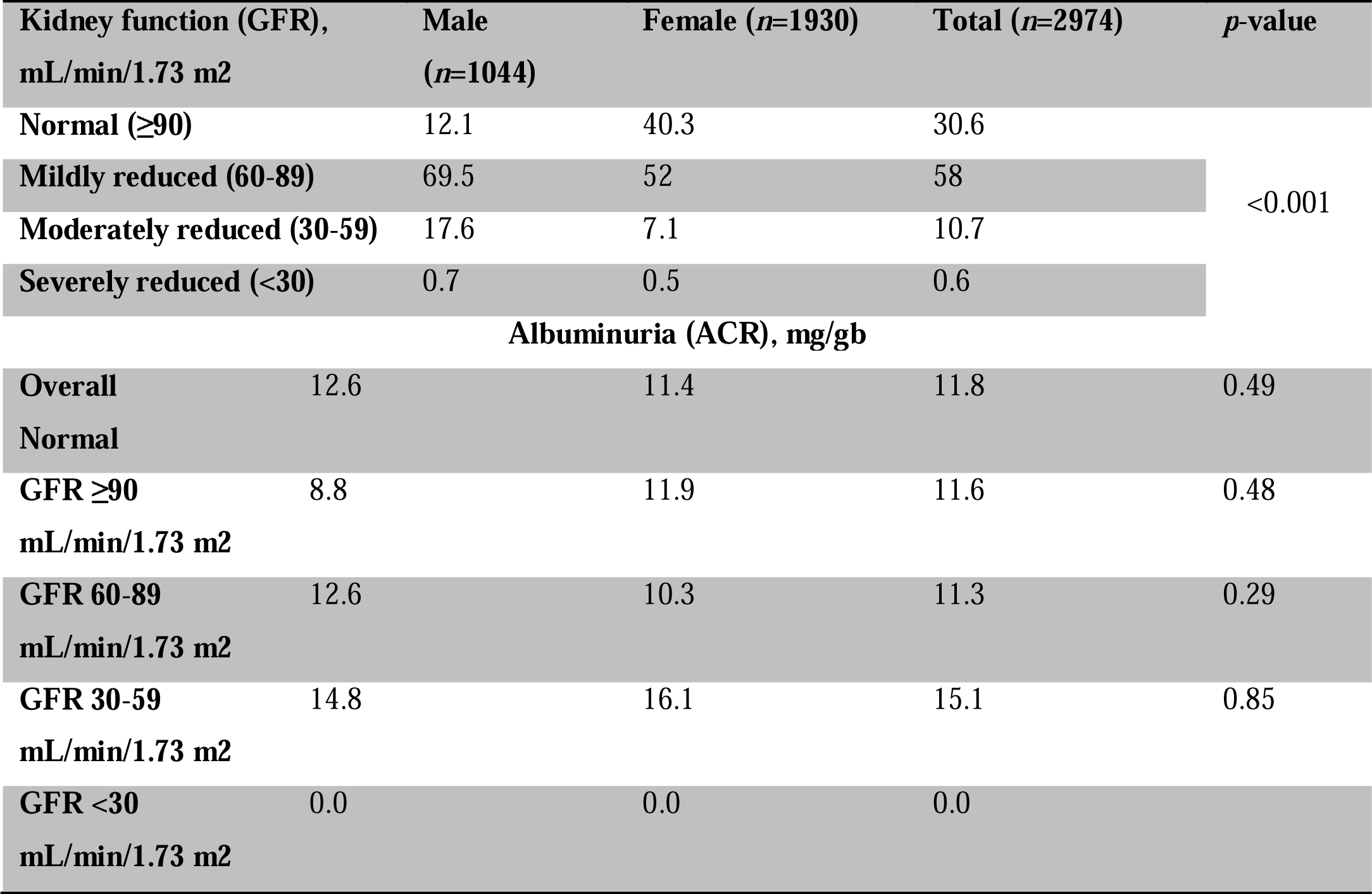
Prevalence of A, Kidney Function and B, Albuminuria categories in the study population.

### Prevalence of chronic kidney disease

In men, 10.8%, 68.3%, 20.2%, 0.5%, 0.2% wasin CKD stages of 1-5, respectively. However, in women, there were radically different distributions of CKD stages: 44.4%, 51.1%, 4.2%, 01% and 0.2% in stages 1-5, respectively (*p*<0.001) (Table 3).

**TABLE 3.** Prevalence of Chronic Kidney Disease (CKD) according to the stages.

| **CKDStage** | **Male (n=1044)** | **Female (n=1930)** | **Total(n=2974)** | **p-value** |
| --- | --- | --- | --- | --- |
| **1** | 10.8 | 44.4 | 32.9 | <0.001 |
| **2** | 68.3 | 51.1 | 57 |  |
| **3** | 20.2 | 4.2 | 9.7 |  |
| **4** | 0.5 | 0.1 | 0.2 |  |
| **5** | 0.2 | 0.2 | 0.2 |  |
| **Total** | 100 | 100 | 100 |  |

### Main risk factors of CKD manifestation

Using univariate and multivariate logistic regression of decreased estimated GFR<60 mL/min/1.73 m2 showed that age (OR=1.06), gender (OR=2.14), diabetes (OR=1.07), hypertension (OR=1.39) and drug use (OR=3.29) were the primary risk factors for CKD in this population, and that BMI was not (Table 4-A). However, univariate and multivariate logistic regression results showed that only age (OR=1.02), diabetes (OR=2.61), and hypertension (OR=1.16) were risk factors for albuminuria (4B).

**TABLE 4.**
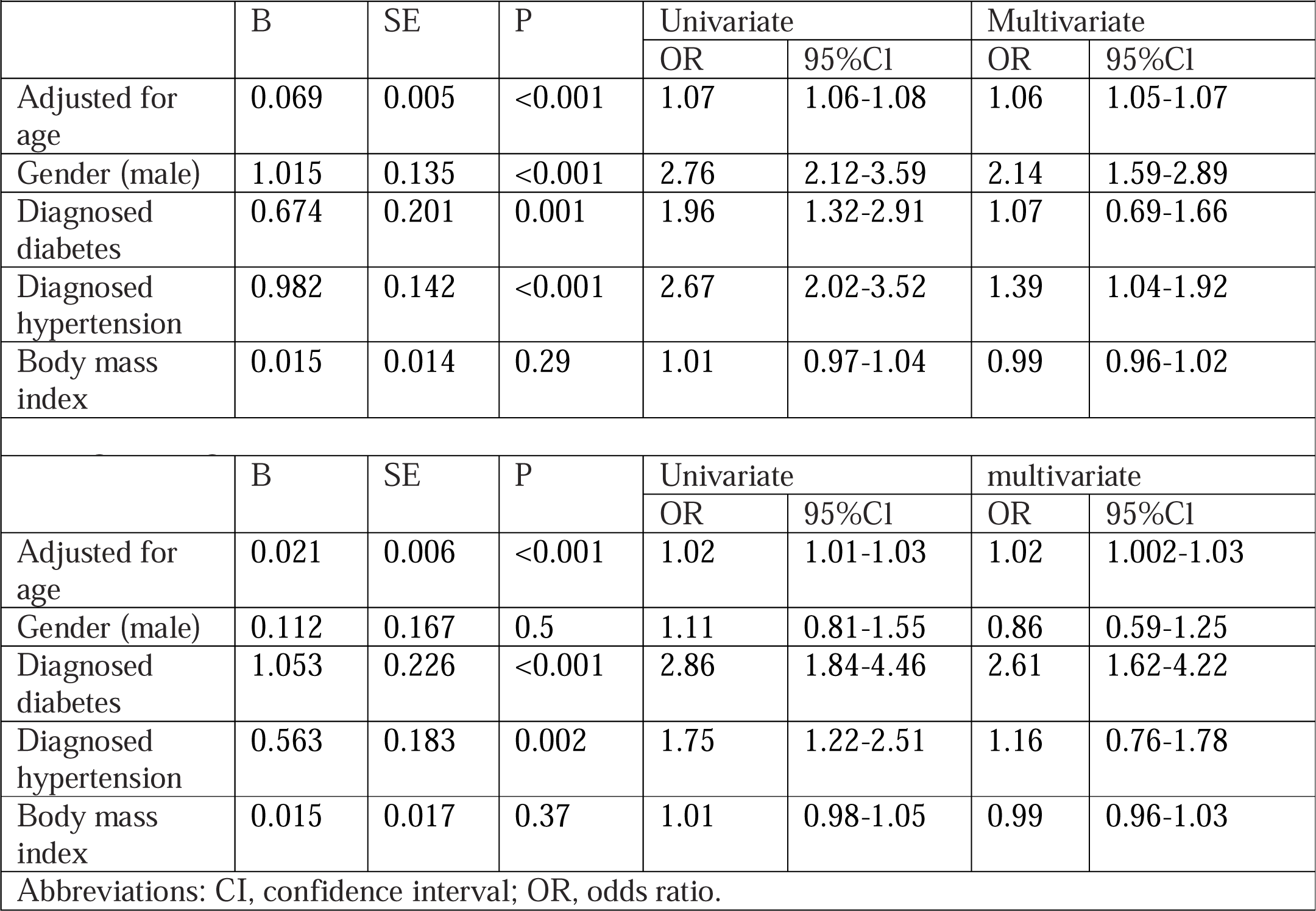
A) Logistic Regression of Decreased Estimated Glomerular Filtration Rate (GFR)<60 mL/min/1.73 m2 in

The distributions of estimated GFR and albuminuria are shown in Fig. 1. It was expected that there would be a normal distribution of GFR and albumin-to-creatinine(Al/Cr) ratio, as GFR had a symmetric normal distribution.However, the Al/Cr ratio was not normal, and most of the subjects (40%)hadratio indices between 5 and 24.99. The prevalence of subjects with GFR below 60 mL/min/1.73 m^2^ was around 12.8%, but most of the subjects (72.6%) had GFR between 60-100 mL/min/1.73 m^2^.

**FIGURE 1.**
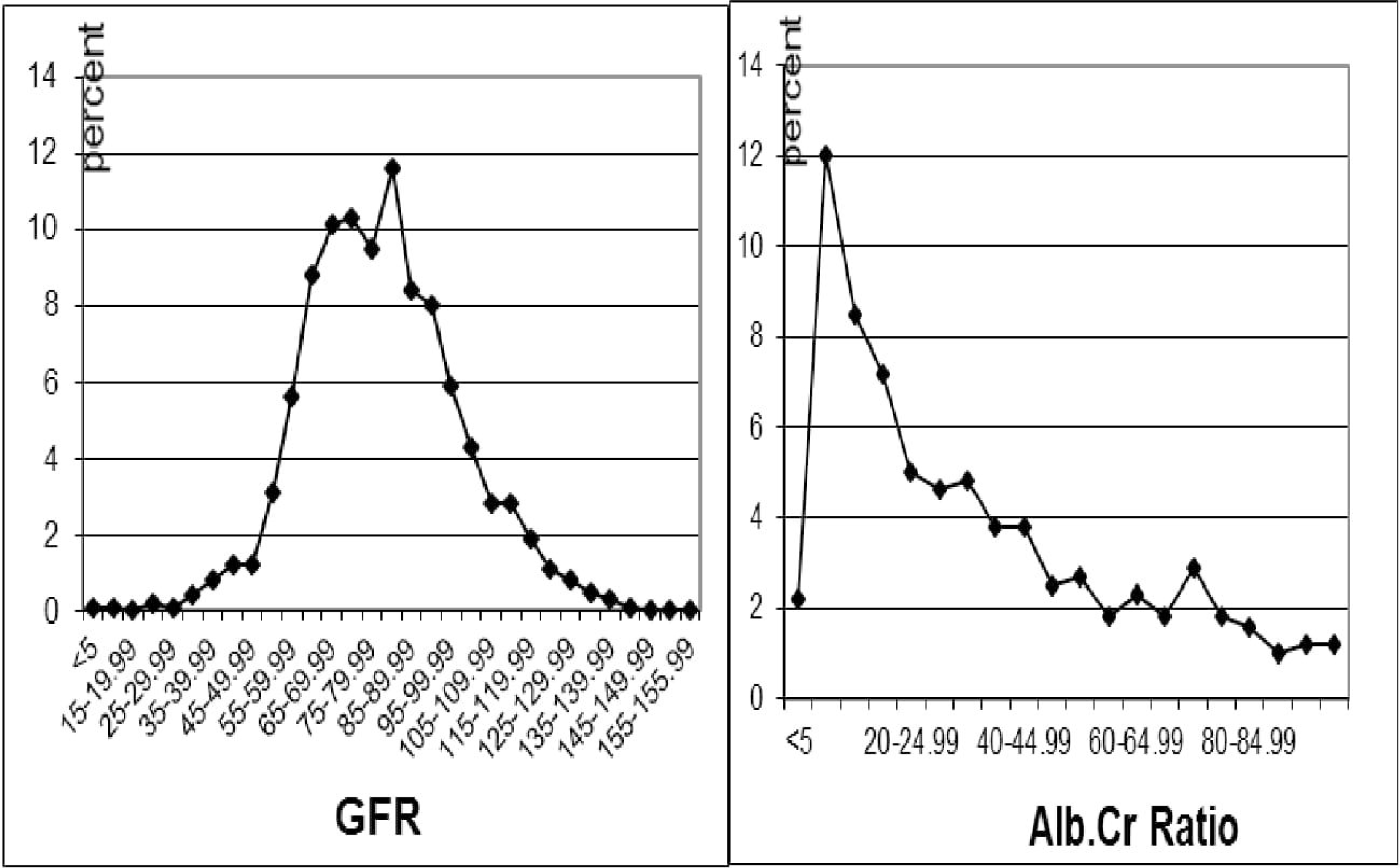
The distribution of estimated **A**, GFR and **B**, albuminuria.

The prevalence of CKD increased with age, as expected. As Fig.2 shows, the under curve area demonstrates the percentage of CKD stages for each age category; for example, the subjects in stage 3 increased from 1.30% in age groups<30 to 3%, 10%, 23.3% and 39.8% in age groups of 30-39, 60-69 and >70, respectively. There were no cases of stages of 4 and 5 in age groups <30, 30-39, 40-59 and 60-69. However,stage 4 and 5 cases were found in 0.30% and 0.40% of participants in age groups 40-59 and in 1% for the age group >70 years. These trends in increasing in CKD stages according to age were expected, as instances of a GFR below 90 mL/min/1.73 m^2^ increases with age.

**FIGURE 2.**
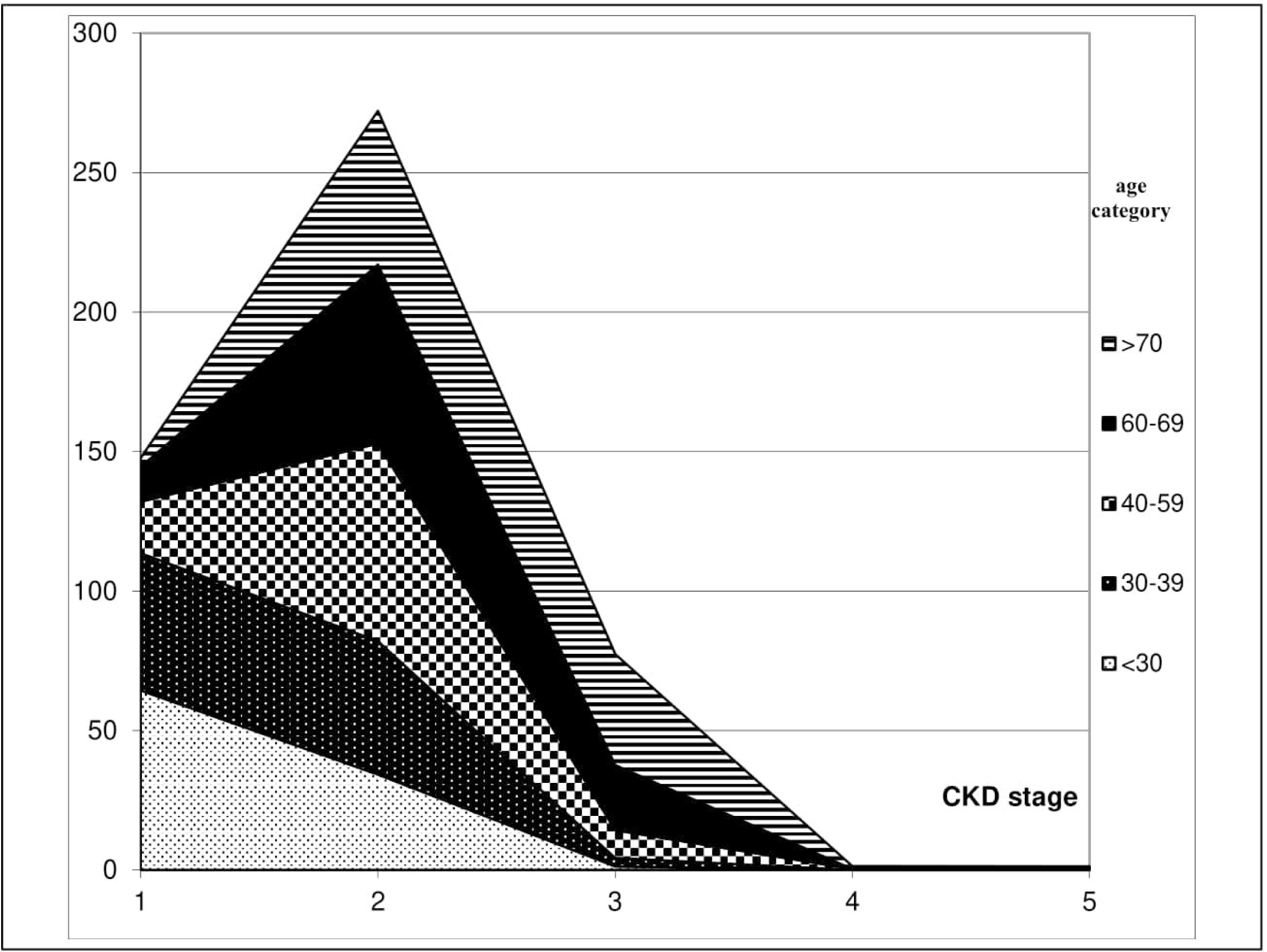
Prevalence of Chronic Kidney Disease (CKD) Stages by Age Group. The under curve area demonstrates the percentage of CKD stages for each age categories.

## DISCUSSION

In this study, the prevalence of CKD was investigated in Mashhad, a city with a population of more than 3 million inhabitants and a moving population of around 20 to 30 million people per year. Mashhad is the second-most-populated city in Iran and is the capital of Razavi Khorasan province. It has a highly-mixed population of Arab, Turkic and Mongolian and Afghan tribes. This diversity makes it a city of interest for an investigation into the main risk factors for CKD.

Although smoking was not a risk factor, addiction was a strong risk factor (OR=3.25) for CKD occurrence. There are two reasons for this.Firstly, opium is the main drug that is popular in Iranian drug culture, and it can cause kidney damage.^15^Our findings in the lab show that most of commercial opiumis contaminated by heavy metals, particularly lead which can intensify tissue injury.^16^ Secondly, many young drug users use amphetamine derivatives. These drugs and their contaminations are more likely the main reasons for renal malfunction in drug users.

The progressive aging of the subjects and the epidemics of obesity and diabetes in many countries complicates to the worldwide burden of CKD.^17^ A cohort study in Iran between 1997 and 2000 demonstrated with the MDRD equation that the overall prevalence of CKD was 18.9% and age-adjusted prevalence was 14.9%.^13^ In our study, the prevalence of CKD in Mashhad was 10% strongly lower than other studies in Iran.A precise scientific evaluation of some of those studies clarifies that their methodology were not adjusted for CKD and at the planning stage not intended to measure its prevalence, thus may have had biases.

This study demonstrated that the age distribution is profound in the age group 40-59 and older, in which CKD seems to be more common; 36% of the subjects were between 40 and 60. This was expected as CKD is more frequent in older subjects (*p*<0.001).CKD affects 10% to 15% of adults in the United States, Europe, and Asia^9,18^, and prevalence increases with age.^9^This implies that low eGFR reflects only aging, and some studies suggest that existing CKD guidelines should be used in older population or subjects.^19-22^

In the present study, hypertension in uni- and multi-variant regression was not a risk factor in our region as a middle class population. CKD is a multi-factor disease with wide variation in risks the world. Growing evidence demonstrated that even diabetes and hypertension in more developed countries associated with CKD, in low and middle income countries apart from race and age, other risk factors such as herbal medications, life style, diet, non-communicable diseases, genetic factors, and water scarcity factors had strong association with kidney failure.^23,24^

On the other hand, in the literature there are studies in favour or opposition of association between hypertension and renal failure, in Italy compared with the USA, diabetes and hypertension largely fail to explain the lower prevalence of CKD. Furthermore, in a review, by Luca De Nicola, these controversial results have been discussed, it is stated that “four countries with lower CKD prevalence (6.8% overall) had a higher (rather than lower) prevalence of hypertension and then the four countries showing higher CKD prevalence (12.2% overall)”.^23^

Therefore, in our region with emphasis on the heterogeneity of the residents, hypertension did not meet significant association with CKD. Taken together, by deep observation on renal failure and staging, our understanding of hypertension and CKD as direct cause radically changed, it is more likely that two factors being possibly attributable to the amelioration of kidney damages including, endothelial dysfunction and inflammation.

The mean BMI of 25-30 (overweight) was more frequent in this study (39.8%), which, using conventional statistical analysis, shows that obesity is a risk factor for CKD (*p*<0.001). However, univariate and multivariate logistic regression showed that BMI was not a risk factor either for decreased GFR or for albuminuria. One study demonstrated that, as BMI decreased, the incidence of ESRD also declined. However, this does not suggest that low BMI impliesa low risk of ESRD.^25^In fact, Ramirez *et al*. demonstrated that patients with very low BMIs had an increased risk of proteinuria.^11^

Bosma*et al*.suggested that the effects of BMI on renal function are not only limited to manifestations of obesity but also other factors, such as diabetes.^26^

The controversial impact of BMI on survival in general and renal failure in particular has been comprehensively reviewed by elsewhere.^27,28^Briefly, in renal failure, body mass and particularly, BMI may be confused by hydration disorders, different stages of CKD and in the elderly. Therefore, to resolve the controversy around BMI in survival and renal failure, it is pivotal to take into account muscle mass, fat or adipose tissue mass, fat distribution and lean body mass rather than BMI. The molecular studies showed that, although, leptin and adiponectin secretion from adipose tissue require for energy and glucose homeostasis, the production of adipokines (TNF-α,and IL-6) promote inflammation which might be complicated in diseases manifestation. Therefor instead of BMI and obesity, the direct mechanisms of adipose tissue on renal failure and survival must take into account.

Multiple studies in Iran demonstrated that female gender was the strongest risk factor for CKD.^13,29^ However, in this study, as table 1 shows, there were associations between GFR and gender, serum creatinine and gender(*p*<0.001).Around 17.6% of males and 7.1% of females had moderately reduced GFR, and 0.7% of males and 0.5% of femaleshad severely reduced GFR. Therefore, the indices for renal function in females are generally better than in males.

In the present study, 10.8%, 68.3%, 20.2%, 0.5% and 0.2% of males were in CKD stages 1-5, respectively. However, in females, there was a radically different distribution of CKD stages: 44.4%, 51.1%, 4.2%, 0.1% and 0.2% for stages 1-5, respectively (*p*<0.001).Although GFR has a symmetric normal distribution (Fig.1), the Al/Cr ratio does not.Most of the subjects (40%) had Al/Cr ratios between ∼ 5 and 24.99, and 72.6% of subjects were fallen between 60 and 100 mL/min/1.73 m2 for the GFR index.However, as previously suggested,differential errors in GFR estimation cannot be denied.^9^

The prevalence of CKD increases with age, as expected (Fig. 2).For example, the subjects in stage 3 increased from 1.30% in the age group <30 to 3%, 10%, 23.3% and 39.8% in age groups 30-39, 60-69 and >70 years, respectively. There were not any case of stages of 4 and 5 in age groups <30, 30-39, 40-59 and 60-69. However,there was a 0.30% occurrence of stage 4 and a 0.40% occurrence of stage 5 in the age group 40-59 and was a 1% occurrence for the age groups >70 years. These trends in increasing CKD stages in accordance with age were expected, as GFRs below 90 mL/min/1.73 m^2^decreases with age.

Finally, there are limitations in this study; we did not consider deeply the socioeconomic condition of household. Using MDRD method, we did not measure serum creatinine and urine albumin in an interval time in negative subjects to reach lowest bias as finding curable acute kidney can effect on our CKD staging. The dietary history and herbal medications which may impact on kidney function did not record.

In conclusion, these findings show that interpretation of the high prevalence of CKD in this population should be considered in the context of the changes in human behaviours, addiction,and diseasetherapy *vs*. prevention, etiology, disease severity and co-occurring diseases.

Albuminuria is the strongest risk factor for CKD progression and increased risk of cardiovascular diseases.^30^ Reduced GFR in chronic kidney disease, particularly in stages 3 and 4,is associated with complications in the cardiovascular system and high costs of treatment in end-stage renal disease. Taken together, more initiatives are needed to reduce the incidence of CKD, such as greater responsibility among health authorities, an emphasis on therapy instead of prevention, and collecting more reliable data to predict complications of reduced GFR, kidney disease progression and cardiovascular manifestations. Therefore, it isnecessary to establish a world scientific program for reducing CKD as an important cause of disease complications and costs for governments. For example, it is pivotal to establish diagnostic guidelines for early stages of CKD that very likely to prevent progression toward (ESRD) and cardiovascular diseases.

In our country, attention to CKD and its preventive programs is very low, particularly in the early stages of CKD, but the cost of treatment, particularly for ESRD, is very high. Therefore, lack of planning for CKD prevention and reliance on treatment is expected to progress ESDR even more dramatically as the population continues to age.

## Data Availability

All of the row data, SPSS file, statistical analysis and the original materials (except the urine samples)
are available.

## Abbreviations

Al/Cr: albumin-to-creatinine ratio
BMI: Body mass index
CG: Cockcroft and Gault
CKD: Chronic kidney disease
CKD-EPI: Chronic Kidney Disease Epidemiology Collaboration
CVD: cardiovascular disease
ESRD: end-stage renal disease
GFR: glomerular filtration rate
KDOQI: Kidney Disease Outcome Quality Initiative
MDRD: Modification of Diet in Renal Disease
RBC: Red blood cells
TC: total cholesterol
TG: triglyceride
WBC: White blood cells

## ACKNOWLEDGEMENTS

The authors have great thank to Mrs ShohrehYaghootkar who sorted all questionnaires, clinical history, laboratory tests and entered the data to the SPSS software. We greatly appreciate to Mrs SamanehAmiri was our great colleague in arrangement of sample collections from 20 Health centres and running laboratory tests in the ACECR-Central Medical Lab.This study was financially supported by the Vice-Chancellor for Research and Technology, Mashhad University of Medical Sciences, Mashhad, Iran, under Grant **[MUMS 87099]**.

## ETHICS APPROVAL AND INFORM CONSENT

All procedures performed in studies involving human participants were in accordance with the ethical standards of the institutional and/or national research committee and with the 1964 Helsinki declaration and its later amendments or comparable ethical standards. This study was reviewed, approved and supervised by the Biomedical Research Ethics Committee of Mashhad University of Medical Sciences **[IR**.**MUMS**.**REC**.**87099]**. Informed consent was obtained from all individual participants included in the study.

## DATA AND MATERIALS AVAILABILITY STATEMENT

The data that support the findings of this study are included in this manuscript and available from the corresponding author **(S**.**A**.**R. Rezaee)**upon reasonable request.

## AUTHOR’S CONTRIBUTIONS

S.A.R. Rezaee and T. Hassannia designed the study and arranged the laboratory tests. M. Khajedaluee did the statistical designs and supervised the data collections. S.A.R. Rezaee and S. Ahmadi Ghezeldasht supervised laboratory tests supervision and quality controls. T. Hassannia and M. Khajahdaluee run the workshops for physicians and questioners. M. Khajedaluee and S. Ahmadi Ghezeldasht performed the statistical analysis of the data. R. Hekmat as nephrologist was one of research advisors, S. Ahmadi Ghezeldasht and A. Mosavat wrote the draft. S.A.R. Rezaee, A. Mosavat and T. Hassannia revised and completed the manuscript.

